# Per capita COVID-19 Case Rates are Lower in U.S. Counties Voting more Heavily Democratic in the 2016 Presidential Election, except not in States with a Republican Governor and Legislature

**DOI:** 10.1101/2020.11.03.20225425

**Authors:** Lloyd Chambless

## Abstract

In our recent paper **Why do per capita COVID-19 Case Rates Differ Between U**.**S. States?** we established that U.S. states with a Democratic governor and a Democratic legislature have lower COVID-19 per capita case rates than states with a Republican governor and a Republican legislature, and case rates of states with a mixed government fall between the two. This difference remained after accounting for differences between states in several demographic and socio-economic variables. In a recent working paper **The Changing Political Geographies of COVID-19 in the U**.**S**. it was found that that early in the pandemic U.S. counties at higher levels of percentage Democratic vote in the 2016 presidential election had higher weekly per capita COVID-19 rates, but that the situation was in the opposite direction by August 2020. We show here that counties with a higher percentage of Democratic vote in the 2016 presidential election have a lower mean cumulative per capita rate of COVID-19 cases and of COVID-19 deaths, adjusted for county demographic and socio-economic characteristics, but only for counties in states that currently have a Democratic governor and both chambers of the legislature Democratic or in states that have a mixed government, but not for states that currently have a Republican governor and both chambers in the legislature Republican. One possible contributor to this difference is that some state Republican governments have restricted local action to fight the spread of COVID-19.

## Introduction

In our recent paper **Why do per capita COVID-19 Case Rates Differ Between U**.**S. States?**^1^ we established that U.S. states with a Democratic governor and a Democratic legislature have lower COVID-19 per capita case rates than states with a Republican governor and a Republican legislature, and case rates of states with a mixed government fall between the two. This difference remained after accounting for differences between states in several demographic and socio-economic variables. In a recent working paper **The Changing Political Geographies of COVID-19 in the U**.**S**.^2^ by Krieger et al, the authors classified U.S. counties in five equal groups according to the percentage of 2016 presidential vote in each county that was Democratic (or that was Republican, or in terms of the difference between the Republican and Democratic percentages – all three ways determine the same groups). They found that early in the pandemic U.S counties in higher categories of percentage Democratic vote had higher weekly per capita COVID-19 rates, and in an approximately graded manner, but that the situation was in the opposite direction by August 2020. Their findings for weekly per capita COVID-19 death rates were similar.

In this report we also analyze per capita COVID-19 case and death rates for the U.S. counties, except we consider the cumulative cases and deaths through October 15, 2020. We adjust the differences in COVID-19 case and death rates between the levels of county Democratic percentage vote in the 2016 election by the same demographic and socio-economic factors considered in our earlier paper. Further, because we already know about the differences in COVID-19 rates between states according to party in power, we first assess whether the association between COVID-19 rates and level of Democratic vote in 2016 differs by current party in power.

## Methods

We refer the reader to our earlier paper for details of our methods, which we only briefly summarize here, and we also mention issues particular to the analysis of county data.

### COVID-19 per capita case and death rates

The counts of COVID-19 cases and deaths through October 15, 2020 are available from the same source as the state data, though data for the five boroughs of New York City come from a separate data set there. The population denominators to define the rates again come from the U.S. Census Bureau. We use the separate. Excluded from our analysis are seven counties in the U.S. Census county list that do not have case counts, five counties in the list of case counts that do not match the Census Bureau list of counties, and one county (Trousdale, TN) that is an extreme outlier in case rate (21,817 per 100,000), leaving 3132 counties in the analysis.

### County demographic and socio-economic characteristics

For percentage of people living in a household with income below the poverty level; median income; percentage of the population living in an urban area; population density (number of people per square mile); percentage of the county population that is Hispanic of any race, and the percentage of the population that is African-American and not Hispanic; median age; percentage of the population above the age of 25 with a bachelor’s degree or higher; and the percentage of the population aged 19-64 that is uninsured, we use the same general Census Bureau sources for counties that we used for states.

### Political status of the counties and states

As in the previous paper we classify states into three categories according to the current party in power: those with governor and both chambers of the state legislature Republican, those with governor and both chambers of the state legislature Democratic, and the others, with mixed government. We also use the same source for county vote totals for the 2016 presidential elections as we used for states, except for Alaska. The state of Alaska does not present the actual presidential vote by county, but estimates of the county totals are elsewhere available.^3^ We group this percentage Democratic of the 2016 presidential vote into four levels. Because of the lopsided number of counties that voted Republican in 2016, we do not use 4 equal groups of percentage Democratic vote, but choose the cutpoints for the lower two groups to assure at least 100 counties in these groups in Democrat party-in-power states.

### Date

As previously we also consider adjusting for the date of first case in each county in the case analysis and the median date of death in each county in the death analysis. For counties without COVID-19 deaths we used the mid date between date of first case and October 15, 2020.

### Analysis

The unit of analysis is U.S. counties. To study the association between vote in the 2016 presidential election and COVID-19 case rate in the counties it is important to control for the county characteristics, in order to assess the level of association that is independent of those characteristics. To this end we apply multiple linear regression of case rate as a function of the county characteristics, the date variable, and the Democratic percentage of the 2016 presidential election vote by county, grouped into four levels. Because of the high correlation between percentage poverty, median income, and percentage with a bachelor’s degree, we replace these latter two variables by their residuals from their prediction as a function of the other county characteristics, including percentage of poverty. Because we have a large sample we are able to include quadratic terms for the county characteristics and the date variable, and we retain terms if they are statistically significant in the overall model or their exclusion changes the estimates of the coefficients of the Democratic vote variables by more than 10%. Since we know from our previous paper that case rate is associated with the party in power in the states, we also include party-in-power variables. We first assess whether the association between case rate and county Democratic percentage of the 2016 vote varies by party in power in the state. We do this by including the relevant interaction terms in the model and then testing the interactions with a six d.f. F-test. Since the interactions were indeed statistically significant, so that the associations of interest do vary by party in power, we then fit separate models for counties belonging to each of the different party-in-power groups, and present results only separately for these three groups. The model directly estimates for each party-in-power group of states the mean differences between the four levels of percentage Democratic vote in the counties for the 2016 presidential election, adjusted for the county characteristics and the date variable. For ease of interpretation we also estimate absolute adjusted means of case rate by percentage Democratic vote, using predicted values from the model, with the adjusting variables set to their means. The analysis of COVID-19 of death rates is similar, since the interaction terms were also statistically significant, so that the results again differ between the three types of states.

We also confirm the results of the case analyses done with ordinary multiple linear regression using a method well-suited to count data, namely a multiplicative (log linear) negative binomial regression model using SAS Proc Genmod. Because there are 556 counties with no COVID-19 deaths we do not take this approach with death rate analysis.

Analysis was done with SAS 9.4, the figures were done with Excel (Microsoft 365).

## Results

Table 1 shows the number of counties cross-classified by level of Democratic vote in the 2016 election and by party in power in the state to which the county belonged. Note that over half of the counties, 1691, are in states with Republican party in power, while only 600 counties are in states with Democratic party in power.

**Table 1:**
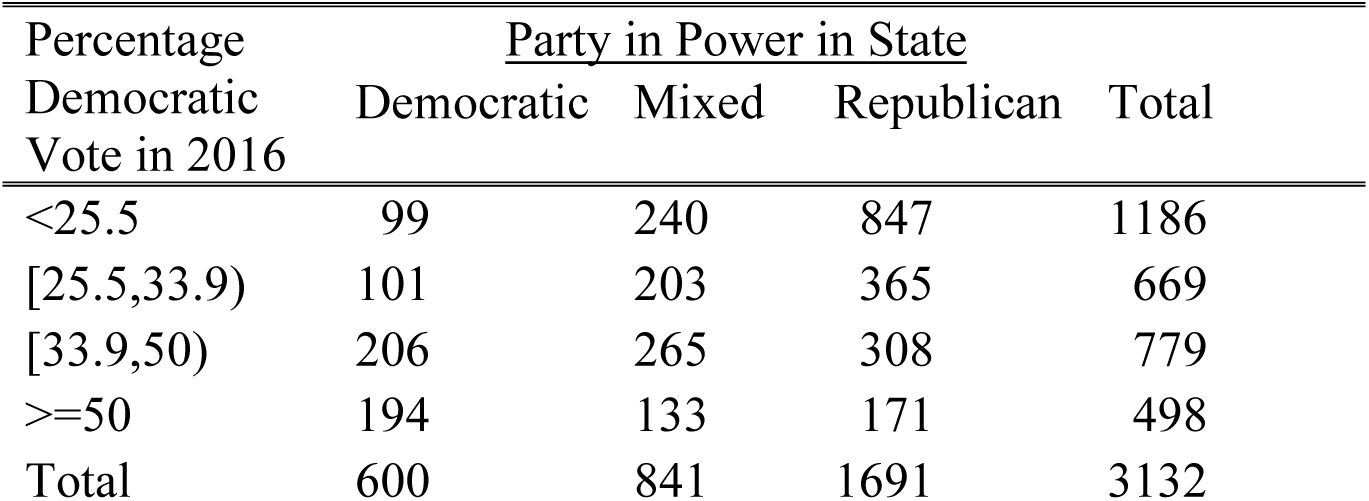
Number of Counties by Percent of Democratic Vote in the 2016 Presidential Election and the Party in Power in the State

The final model for case rate included percentage Hispanic, population density, percentage urban, percentage with bachelor’s degree (residual) and the date variable as linear terms, and included percentage Black, median age, percentage uninsured, percentage in poverty, and median income (residual) as linear and quadratic terms. The model for all 3132 counties also included the variables for the four-level percentage Democratic 2016 county vote, the three-level party-in-power variables, and the interaction of these variables. This model had R^2^ = 0.35 and had statistically significant interactions (p < 0.0001), showing that the associations between case rate and percentage Democratic 2016 county vote differed by state party in power. Thus, our final models including the variables for the four-level percentage Democratic 2016 county vote were fit separately by state party in power. Table 2 shows for each party-in-power group of states the mean case rate differences between counties at each level of percentage Democratic vote in 2016 and those at the lowest level, adjusted for the other variables in the model, together with the p-values to assess statistical significance (p < 0.05). For states with Democratic party in power there is a strong graded difference in adjusted mean case rates, with counties having a higher percentage Democratic vote in 2016 having a lower rate of COVID-19 cases per 100,000 people. For states with a mixed government there was also a graded association, though less strong than with counties in Democratic-led states. For states with the Republican Party in power there was no clear difference between counties at the four levels of percentage Democratic vote.

**Table 2:** Adjusted Difference in Mean Cases per 100,000 Between the Top Three Levels of Percentage of Democratic County Vote in the 2016 Presidential Election and the Bottom Level (p-values in parentheses)

| State Party in Power | Percentage of Democratic Vote |  |  |
| --- | --- | --- | --- |
| | [25.5,33.9) | [33.9,50) | $\geq 50$ |
| Democratic | -289 (0.072) | -547 (< 0.001) | -626 (0.001) |
| Mixed | -31 (0.772) | -106 (0.346) | -580 (< 0.001) |
| Republican | 168 (0.072) | -15 (0.893) | 17 (0.931) |

An alternative presentation of the same results except showing absolute adjusted mean case rates instead of adjusted differences between levels of Democratic county vote is in Figure 1. The figure confirms the findings of the previous paper, now at each level of the percentage Democratic vote of 2016, that there is a graded decrease of cases per 100,000 people as one moves from Republican to Mixed to Democratic-led states. The figure also illustrates the new finding in this paper, that for two of the groups of states, Democratic (blue) and Mixed (purple), as the county percentage Democratic vote in 2016 increases, the adjusted case rate decreases. This is not true for the Republican states (red).

**Figure 1:**
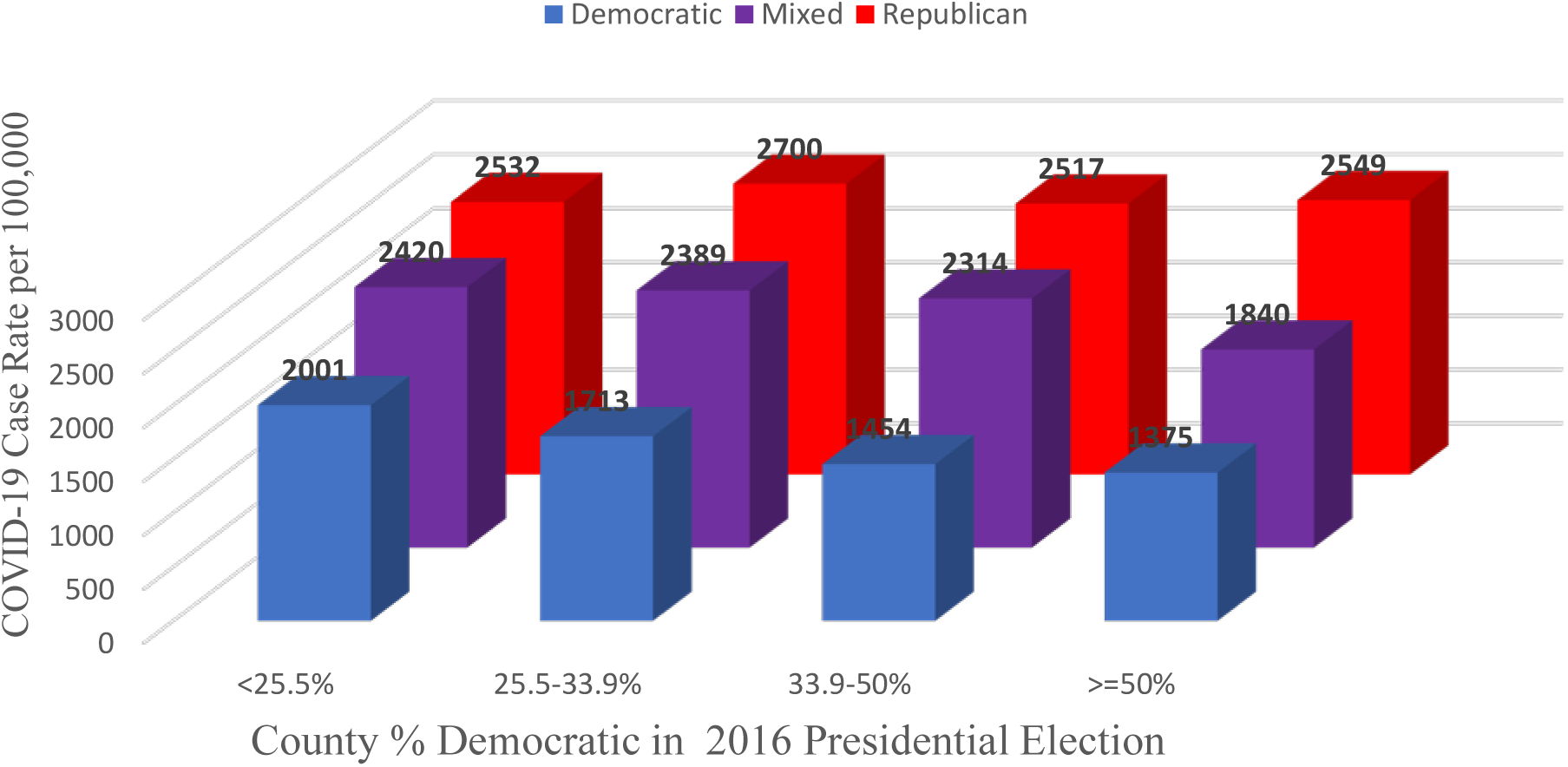
Adjusted Mean COVID-19 Case Rate by Level of Democratic Vote in 2016 Presidential Election and State Party in Power

In the negative binomial modeling of case rate using the same variables as in the linear model, the interaction model for all 3132 counties had R^2^ = 0.32 (after transforming back to the original scale) and again had significant interaction terms (p < 0.0001). In the separate models of the three state party-in-power groups of counties, the results were similar though somewhat stronger than for the linear model. For example, the highest level of four 2016 percentage Democratic vote groups had 54% (p <0.001), 69% (p < 0.001), and 101% (p = 0.854) of the adjusted case rates as did the lowest level, for the Democratic, Mixed, and Republican state party-in-power groups, respectively. Calculating ratios from the rates given from the linear model in Figure 1 we get similar percentages, 69% (p = 0.001), 76% (p < 0.001), and 101% (p = 0.931), respectively.

The final model for death rate included percentage urban, percentage with bachelor’s degree (residual), and median income (residual) as linear terms, and included percentage Hispanic, percentage Black, population density, median age, percentage uninsured, percentage in poverty, and the date variable as linear and quadratic terms. The interaction model for all 3132 counties for death rate had R^2^ = 0.34 and also had statistically significant interactions (p < 0.0001), again showing that the associations between death rate and percentage Democratic 2016 county vote differed by party in power, so again our final models including the variables for the four level percentage Democratic 2016 county vote were fit separately by type of state party in power. Table 3 shows for each party-in-power group of states the adjusted mean death rate differences between counties at each level of percentage Democratic vote in 2016 and those at the lowest level, together with the p-values to assess statistical significance. For states with Democratic party in power, counties in the higher two levels of percentage Democratic vote in 2016 have a lower adjusted mean rate of COVID-19 deaths per 100,000. For states with a Mixed government there was a stronger graded association – the higher the counties’ percentage Democratic vote in 2016, the lower the adjusted COVID-19 death rate. For Republican-led states there was a suggestion of an association in the opposite direction, though not statistically significant.

**Table 3:** Adjusted Difference in Mean Deaths per 100,000 Between the Top Three Levels of Percentage of Democratic County Vote in the 2016 Presidential Election and the Bottom Level (p-values in parentheses)

| State Party in Power | Percentage of Democratic Vote |  |  |
| --- | --- | --- | --- |
| | [25.5,33.9) | [33.9,50) | $\geq 50$ |
| Democratic | 1.6 (0.812) | -12.1 (0.057) | -6.0 (0.450) |
| Mixed | -8.6 (0.021) | -11.2 (0.005) | -14.1 (0.015) |
| Republican | 6.1 (0.053) | 3.8 (0.333) | 0.7 (0.917) |

The alternative presentation of these results in terms of absolute adjusted mean death rates instead of differences between levels of Democratic county vote is in Figure 2. The decrease in adjusted COVID-19 death rates in counties with increasing percent Democratic vote in the 2016 presidential election is clear in the Mixed states (purple), less pronounced in Democratic-led states (blue), and absent in Republican-led states.

**Figure 2:**
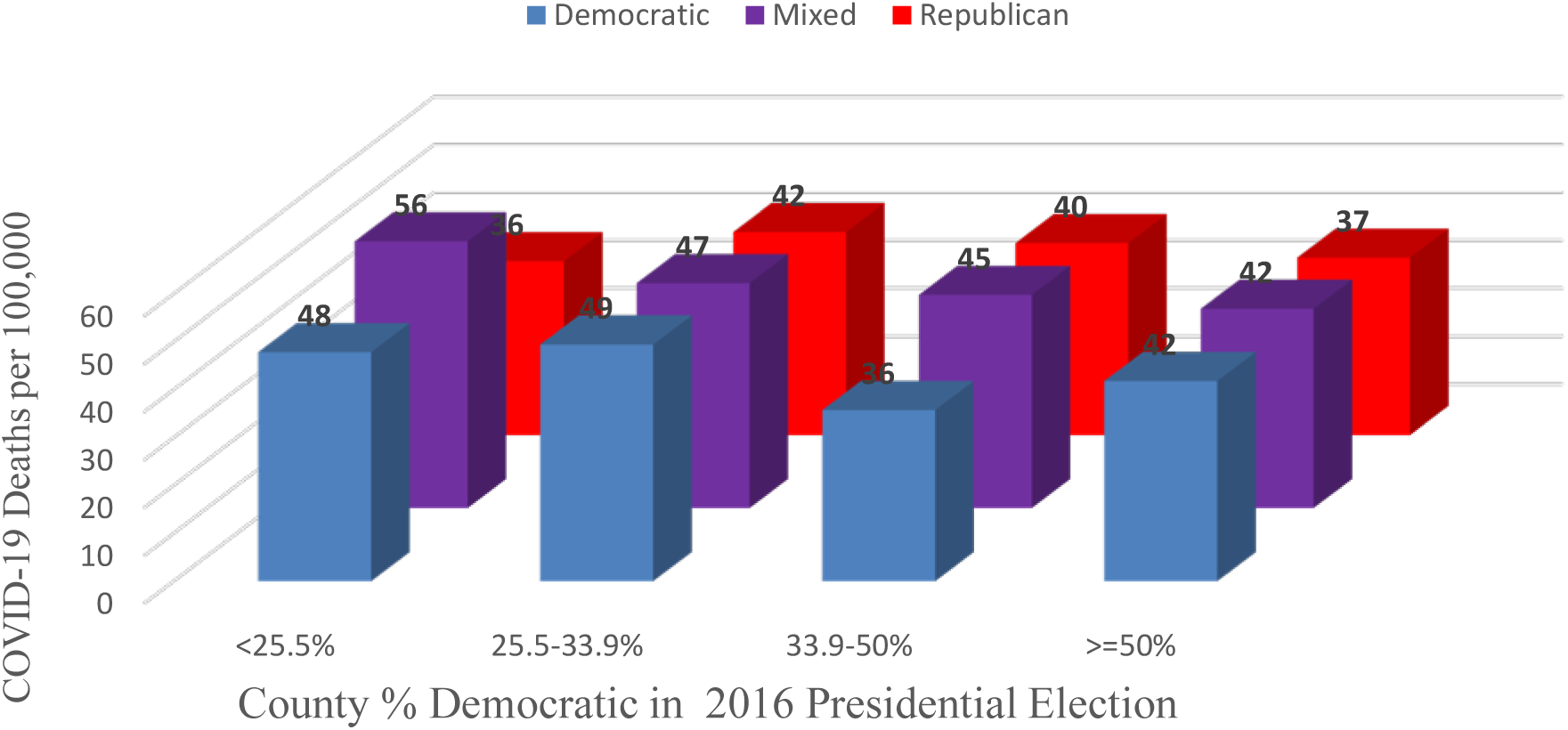
Adjusted Mean COVID-19 Death Rate by Level of Democratic Vote in 2016 Presidential Election and State Party in Power

## Discussion

We have shown that counties with a higher percentage of Democratic vote in the 2016 presidential election have a lower mean cumulative per capita rate of COVID-19 cases and of COVID-19 deaths, adjusted for county demographic and socio-economic characteristics and date variables, but only for counties in states that currently have a Democratic governor and both chambers of the legislature Democratic or in states that have a mixed government, but not for states that currently have a Republican governor and both chambers in the legislature Republican. One possible contributor to this difference is that some state Republican governments have restricted local action to fight the spread of COVID-19.^4^

## Data Availability

All data is publicly available on the internet using the 3 links just below and those listed under Data Availability Links
https://github.com/nytimes/Covid-19-data
https://ballotpedia.org/State_government_trifectas
https://doi.org/10.7910/DVN/42MVDX

https://data.census.gov/cedsci/table?q=race%20by%20state&g=0100000US&tid=ACSDP1Y2018.DP05&hidePreview=true

https://www.census.gov/search-results.html?searchType=web&cssp=SERP&q=median%20household%20income%20by%20state

https://www.census.gov/programs-surveys/geography/guidance/geo-areas/urban-rural/2010-urban-rural.html

https://data.census.gov/cedsci/table?q=race%20by%20state&g=0100000US&tid=ACSDP1Y2018.DP05&hidePreview=true

https://data.census.gov/cedsci/table?q=median%20age%20by%20state&g=0100000US&tid=ACSST1Y2018.S0101&hidePreview=false

https://data.census.gov/cedsci/table?q=ACSST1Y2019.S1501&tid=ACSST1Y2019.S1501&hidePreview=true

https://data.census.gov/cedsci/table?q=uninsured%20by%20state&g=0100000US.04000.001&tid=ACSST1Y2018.S2701&hidePreview=true

https://rrhelections.com/index.php/2018/02/02/alaska-results-by-county-equivalent-1960-2016/

https://doi.org/10.7910/DVN/IG0UN2

